# Modifiable service-delivery factors, not geography, drive patient satisfaction in rural Sierra Leone: a district-comparative cross-sectional household survey of 679 facility users

**DOI:** 10.64898/2026.08.04.26359754

**Authors:** Alhassan Mayei, Yvonne Schoenemann, Nina Siegert, Lisa Seidelmann, Baillah Molleh, Sulaiman Lakoh, Osman Sankoh

## Abstract

**Background:** Patient-reported satisfaction is a core tracer of health-system responsiveness in Universal Health Coverage (UHC) monitoring, yet its determinants in rural Sierra Leone are poorly characterised. We quantified overall and domain-specific satisfaction and identified modifiable predictors across three rural border districts.

**Methods:** We conducted a cross-sectional, population-based household survey in October 2024 in Kailahun, Kambia and Pujehun districts, using a two-stage cluster design (chiefdoms sampled with probability proportional to size; households sampled at random). A validated 29-item instrument measured overall satisfaction and eight patient-experience domains on five-point Likert scales. The primary outcome was the five-level single-item overall satisfaction rating. We fitted a multivariable proportional-odds ordinal logistic regression, with 95% confidence intervals (CIs) obtained by a cluster bootstrap resampling the 20 chiefdom clusters. Robustness was assessed with binary and composite-outcome sensitivity models.

**Results:** Of 750 respondents, 679 (90.5%) had used a formal health facility in the previous 12 months and formed the analytic sample (510 [75.1%] female; mean age 28.2 years [SD 13.8]). The instrument showed high internal consistency (Cronbach α = 0.87 for the five core domains). Overall, 375/679 (55.2%) were satisfied or very satisfied, ranging from 185/244 (75.8%) in Kambia to 110/224 (49.1%) in Kailahun and 80/211 (37.9%) in Pujehun (χ² = 70.8; p<0.001). In the adjusted model, staff attitude was the strongest predictor of higher satisfaction (adjusted odds ratio [AOR] 2.74, 95% CI 2.03–3.70; p<0.001 per one-point increase), followed by waiting-time satisfaction (AOR 1.89, 1.39–2.46) and medicine availability (AOR 1.43, 1.16–2.10). Travel-time category, facility type and sex were not independently associated. Large district disparities persisted after adjustment: relative to Kambia, the AOR for higher satisfaction was 0.27 (0.15– 0.50) in Kailahun and 0.33 (0.20–0.66) in Pujehun. Adjusted probabilities of high satisfaction were 0.72, 0.48 and 0.43, respectively.

**Conclusions:** Respectful provider behaviour, shorter waits and reliable medicine supply, all amenable to district-level management, were the dominant and actionable drivers of patient satisfaction, whereas geographic distance was not. Persistent between-district gaps call for tailored quality-improvement in Kailahun and Pujehun. Institutionalising routine patient-experience measurement would strengthen accountability for people-centred care and support equitable progress towards UHC.

**Author summary:** Whether people are satisfied with the care they receive shapes whether they return, follow advice and trust the health system, and it is a recognised marker of quality within Universal Health Coverage. Evidence on what drives satisfaction outside Sierra Leone’s capital has been thin, which limits the design of practical improvements in the rural districts where needs are greatest. We surveyed 679 recent health-facility users across three rural border districts and analysed which factors were most strongly associated with higher satisfaction, accounting for the clustered survey design. We found that how patients were treated by staff, how long they waited, and whether medicines were available mattered most, and that all three are within the control of facility and district managers. In contrast, how far patients travelled to reach care did not independently affect satisfaction once they arrived. Satisfaction differed sharply between districts even after adjustment, with Kailahun and Pujehun lagging well behind Kambia, pointing to differences in service delivery rather than in the people served. These findings identify concrete, low-cost priorities, staff communication, queue management and supply reliability, and make the case for routine measurement of patient experience to hold services accountable for people-centred care.

## Introduction

Patient-reported satisfaction is a recognised tracer of health-system responsiveness within the World Health Organization (WHO) framework for monitoring Universal Health Coverage (UHC) and a determinant of continued care-seeking, treatment adherence and trust in services [1,2]. Pooled evidence from sub-Saharan Africa suggests that only about half of facility users report high satisfaction, with long waiting times, poor provider communication and frequent medicine stock-outs the most consistently cited sources of dissatisfaction [3]. Sierra Leone’s National Health Sector Strategic Plan 2021–2025 and its UHC Roadmap 2021–2030 both place people-centred quality at the core of the reform agenda, yet empirical evidence on the determinants of satisfaction beyond the capital remains scarce [2,3].

This evidence gap is consequential in the border districts of Kailahun, Kambia and Pujehun, which together are home to more than one million people and are characterised by difficult terrain, fragile supply chains and recurrent epidemic threats along the frontiers with Guinea and Liberia. These districts have not previously been subject to a systematic, comparative assessment of patient experience. The Health System Strengthening and Epidemic Prevention (HSSEP-III) programme, funded by the German Federal Ministry for Economic Cooperation and Development (BMZ) and implemented by jointly the MoH-SL and GIZ, works in these districts to reinforce routine care and epidemic preparedness through governance, infrastructure and workforce strengthening [4,5]. Embedding a standardised household survey within the programme created an opportunity to generate district-level evidence on community experience of care that is directly relevant to both operational steering and national policy.

We therefore set out to provide a district-comparative, gender-disaggregated analysis of patient satisfaction in rural Sierra Leone. Specifically, we (i) quantified overall and domain-specific satisfaction with primary– and secondary-level services; (ii) estimated the independent associations of facility type, sex, age, visit frequency, travel time, and three modifiable service-experience factors (staff attitude, waiting time and medicine availability) with overall satisfaction; and (iii) compared adjusted satisfaction across districts. By separating modifiable service-delivery factors from fixed contextual characteristics, we aimed to identify concrete, district-level levers for quality improvement and to inform Sierra Leone’s pursuit of equitable UHC.

## Methods

### Study design and setting

We conducted a cross-sectional, population-based household survey in October 2024 in three rural border districts of Sierra Leone: Kailahun (Eastern Province), Kambia (North-West Province) and Pujehun (Southern Province). The districts were purposively selected by the HSSEP programme because of high maternal-mortality burden and elevated vulnerability to cross-border epidemic threats. Each district contains one secondary-level district hospital and a network of community health centres (CHCs) and community health posts, several of which provide basic emergency obstetric and neonatal care. The study is reported in accordance with the STROBE guideline for cross-sectional studies [12] (S1 Checklist).

### Participants and sampling

Eligible respondents were household members of any age who had used a formal health service within the previous 12 months. One eligible respondent was interviewed per household. A two-stage cluster design was used: in the first stage, chiefdoms (primary sampling units) were stratified by district and selected with probability proportional to population size to ensure geographic representativeness; in the second stage, households were drawn at random from updated community lists within selected chiefdoms. Twenty chiefdoms were sampled across the three districts.

The target sample of 750 households (250 per district) was calculated to detect a 10-percentage-point difference in the prevalence of high satisfaction between districts at 95% confidence and 80% power, assuming a design effect of 1.5 and 5% non-response (S2 Text). Minors (<18 years) were eligible where a parent or guardian gave permission and the minor assented; interviews with minors were conducted privately, without facility staff present, and were limited to recent service use and experience of care.

### Data collection and instrument

Six trained enumerators completed a five-day course covering informed consent, digital data capture on Android tablets (KoBoCollect v2023.3) and offline synchronisation. The questionnaire was adapted from a validated patient-experience instrument for low– and middle-income settings and comprised 29 Likert-scale items spanning overall satisfaction and eight service-experience domains (staff attitude, waiting time, cleanliness, medicine availability, perceived quality, communication, cost and feedback), together with sociodemographic and service-use variables. Built-in skip logic and daily supervisory checks minimised entry errors, and encrypted records were uploaded to a secure server when connectivity allowed. Internal consistency of the multi-item scale was high (Cronbach α = 0.87 across the five core service domains; α = 0.92 across all eight domains).

### Variables and outcome

The primary outcome was the single-item overall satisfaction rating with care received, measured on a five-point Likert scale (1 = very dissatisfied to 5 = very satisfied) and analysed as an ordinal variable. For descriptive purposes it was also dichotomised as high satisfaction (score ≥4, i.e. satisfied or very satisfied). Candidate predictors, specified a priori, were district, facility type most recently attended (hospital vs CHC), sex, age (per 10 years), visit frequency in the previous year, travel-time category, and three modifiable service-experience factors treated as continuous per-point scores: staff attitude, waiting-time satisfaction and medicine availability.

### Statistical analysis

Respondent characteristics were summarised overall and by district using frequencies and percentages for categorical variables and means (SD) or medians (IQR) for continuous variables; between-district differences in high satisfaction were tested with the χ² test. The primary model was a multivariable proportional-odds ordinal logistic regression relating the five-level overall satisfaction outcome to the pre-specified predictors. Because the sampling design clustered respondents within 20 chiefdoms, and the number of clusters was modest, 95% CIs and p-values were derived from a non-parametric cluster bootstrap (1,000 resamples of chiefdoms with replacement, refitting the full model in each replicate); this avoids reliance on large-cluster asymptotics. The proportional-odds assumption was examined by comparing coefficients from binary logistic models fitted at each cut-point of the outcome. Adjusted odds ratios (AOR) above 1 indicate higher odds of reporting a higher satisfaction category. To aid interpretation, we computed the model-predicted probability of high satisfaction (score ≥4) for each district, averaging individual predictions and bootstrapping the district means.

Two sensitivity analyses assessed robustness: (i) a binary logistic model for high satisfaction (score ≥4) with the same predictors and cluster-bootstrap CIs; and (ii) an ordinal model using a five-domain composite score in place of the single item. Missingness was minimal: all 679 analytic respondents had complete data on the outcome and service-experience items, so complete-case analysis was used (three respondents whose most-visited facility was neither a hospital nor a CHC were excluded from the regression, leaving n = 676). Analyses were conducted in Python 3.10 (NumPy); analytic code is provided in S3 Code.

### Ethics

Ethical approval for the secondary analysis of the 2024 survey data was granted by the Sierra Leone Ethics and Scientific Review Committee (SLESRC 002/10/2025). The study followed the Declaration of Helsinki and applicable national requirements. All participants provided informed consent before the original survey; written consent was obtained from adults, and written parental/guardian consent plus minor assent for participants under 18 years. Participation was voluntary and all data were de-identified before analysis.

## Results

### Participants

Of 750 respondents who consented and completed the interview, 679 (90.5%) had used a formal health facility in the previous 12 months and were included in the analysis (Fig 1): 224 in Kailahun, 244 in Kambia and 211 in Pujehun. Among the 679 facility users, 510 (75.1%) were female and the mean age was 28.2 years (SD 13.8; median 23, IQR 19–35). Most were aged 18– 24 years (272; 40.1%), married (465; 68.5%), and had either no formal education (254; 37.4%) or secondary education (257; 37.8%). CHCs were the most frequently used facility type (443; 65.2%), followed by government hospitals (233; 34.3%). The commonest reasons for the most recent visit were treatment (298; 43.9%), child-health services (134; 19.7%) and maternal-health services (120; 17.7%). Respondent characteristics differed appreciably across districts, including a higher share of hospital use and of older respondents in Kambia and a younger, predominantly female profile in Pujehun (Table 1).

**Fig 1.**
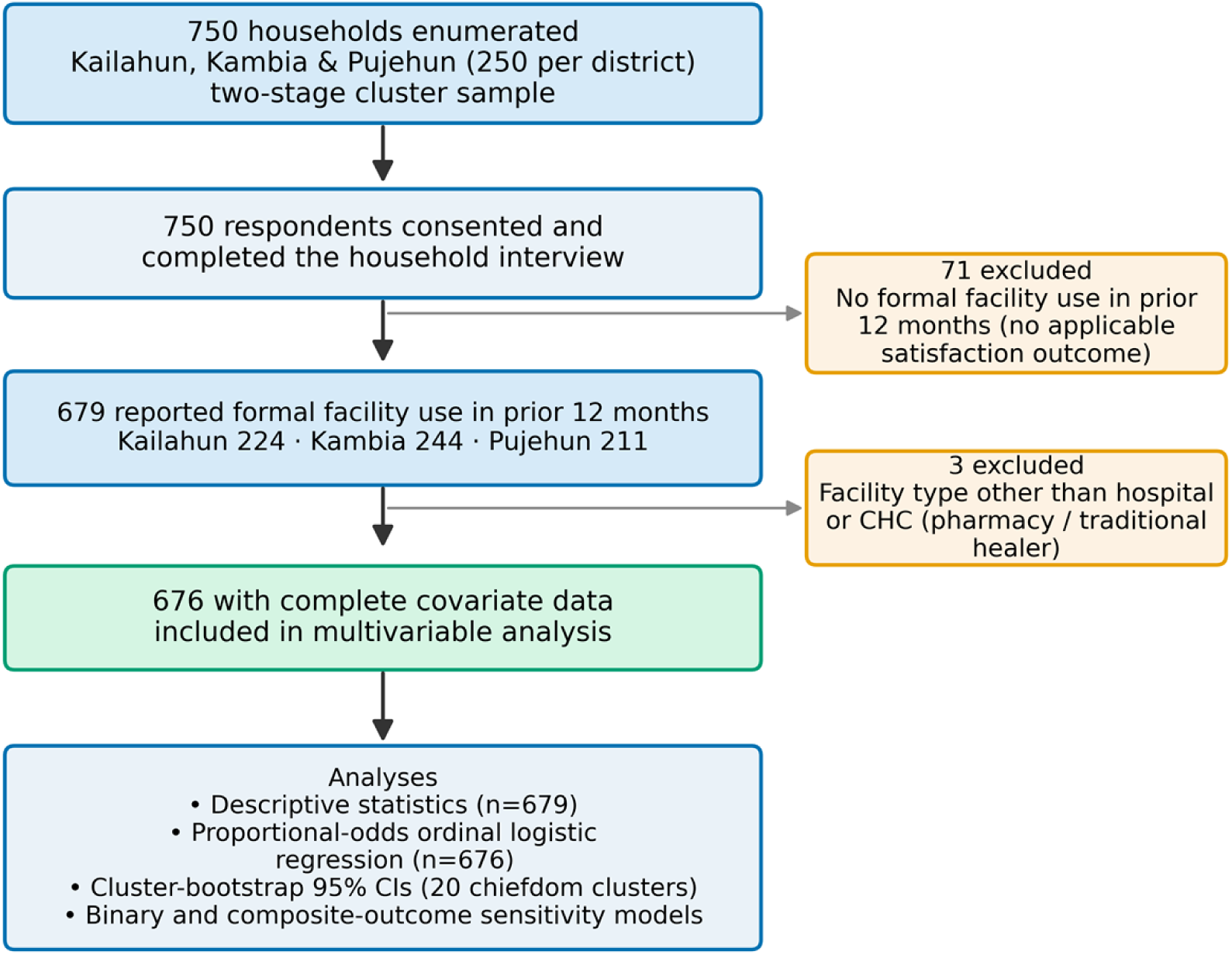
Participant flow. Flow of respondents from household enumeration to the analytic and regression samples, with reasons for exclusion.

**Table 1.** Characteristics of respondents reporting formal health-facility use in the previous 12 months, overall and by district (n = 679)

| Characteristic | Overall (n=679) | Kailahun (n=224) | Kambia (n=244) | Pujehun (n=211) |
| --- | --- | --- | --- | --- |
| **Age** |  |  |  |  |
| Age, mean (SD) | 28.2 (13.8) | 27.6 (13.4) | 33.5 (16.7) | 22.7 (6.3) |
| Age, median (IQR) | 23 (19-35) | 22 (19-35) | 30 (19-43) | 20 (19-24) |
| <18 | 99 (14.6) | 29 (12.9) | 48 (19.7) | 22 (10.4) |
| 18-24 | 272 (40.1) | 100 (44.6) | 36 (14.8) | 136 (64.5) |
| 25-34 | 136 (20.0) | 37 (16.5) | 62 (25.4) | 37 (17.5) |
| 35-44 | 88 (13.0) | 34 (15.2) | 39 (16.0) | 15 (7.1) |
| 45+ | 84 (12.4) | 24 (10.7) | 59 (24.2) | 1 (0.5) |
| **Sex, n (%)** |  |  |  |  |
| Female | 510 (75.1) | 170 (75.9) | 150 (61.5) | 190 (90.0) |
| Male | 169 (24.9) | 54 (24.1) | 94 (38.5) | 21 (10.0) |
| **Marital status, n (%)** |  |  |  |  |
| Married | 465 (68.5) | 165 (73.7) | 176 (72.1) | 124 (58.8) |
| Single | 173 (25.5) | 47 (21.0) | 51 (20.9) | 75 (35.5) |
| Widowed | 34 (5.0) | 10 (4.5) | 15 (6.1) | 9 (4.3) |
| Separated | 5 (0.7) | 1 (0.4) | 2 (0.8) | 2 (0.9) |
| Divorced | 2 (0.3) | 1 (0.4) | 0 (0.0) | 1 (0.5) |
| **Education, n (%)** |  |  |  |  |
| No formal education | 254 (37.4) | 77 (34.4) | 113 (46.3) | 64 (30.3) |
| Primary | 132 (19.4) | 44 (19.6) | 54 (22.1) | 34 (16.1) |
| **Secondary** | 257 (37.8) | 93 (41.5) | 54 (22.1) | 110 (52.1) |
| **Tertiary** | 15 (2.2) | 9 (4.0) | 6 (2.5) | 0 (0.0) |
| **Other** | 21 (3.1) | 1 (0.4) | 17 (7.0) | 3 (1.4) |
| **Facility type, n (%)** |  |  |  |  |
| **Facility: CHC** | 443 (65.2) | 161 (71.9) | 105 (43.0) | 177 (83.9) |
| **Facility: Hospital** | 233 (34.3) | 61 (27.2) | 138 (56.6) | 34 (16.1) |
| **Reason for most recent visit, n (%)** |  |  |  |  |
| **Treatment** | 298 (43.9) | 103 (46.0) | 137 (56.1) | 58 (27.5) |
| **Child health** | 134 (19.7) | 38 (17.0) | 54 (22.1) | 42 (19.9) |
| **Maternal health** | 120 (17.7) | 33 (14.7) | 47 (19.3) | 40 (19.0) |
| **Immunization** | 62 (9.1) | 24 (10.7) | 2 (0.8) | 36 (17.1) |
| **Family planning** | 38 (5.6) | 7 (3.1) | 2 (0.8) | 29 (13.7) |
| **General checkup** | 27 (4.0) | 19 (8.5) | 2 (0.8) | 6 (2.8) |
| **Service use** |  |  |  |  |
| **Visit freq, median (IQR)** | 3 (2-5) | 3 (2-5) | 3 (2-5) | 3 (2-5) |
| **Travel time to facility, n (%)** |  |  |  |  |
| **<30 min** | 122 (18.0) | 49 (21.9) | 22 (9.0) | 51 (24.2) |
| **30 min-1 h** | 263 (38.7) | 74 (33.0) | 91 (37.3) | 98 (46.4) |
| **1-2 h** | 211 (31.1) | 80 (35.7) | 83 (34.0) | 48 (22.7) |
| **>2 h** | 83 (12.2) | 21 (9.4) | 48 (19.7) | 14 (6.6) |
| **Walking to facility** | 478 (70.4) | 166 (74.1) | 123 (50.4) | 189 (89.6) |
| **Outcome** |  |  |  |  |
| **High satisfaction (≥4)** | 375 (55.2) | 110 (49.1) | 185 (75.8) | 80 (37.9) |

*SD = standard deviation; IQR = interquartile range; CHC = community health centre. Percentages are column percentages within each district and may not sum to 100 owing to rounding. High satisfaction = overall rating of 4 or 5 on a five-point scale.*

### Overall and domain-specific satisfaction

Overall, 375 of 679 respondents (55.2%) were satisfied or very satisfied with the care received. High satisfaction varied markedly by district: 185/244 (75.8%) in Kambia, 110/224 (49.1%) in Kailahun and 80/211 (37.9%) in Pujehun (χ² = 70.8, df = 2; p<0.001) (Fig 2A). Domain-level results showed the same district ordering and pinpointed the weakest links in the service experience: satisfaction with medicine availability (33.7% overall) and cost (34.3%) was lowest across all districts, while cleanliness (74.4%) and communication (68.0%) were highest (Fig 2B). Pujehun recorded the lowest satisfaction on six of eight domains, most strikingly for staff attitude (39.3% vs 74.2% in Kambia) and medicine availability (17.5% vs 46.3%).

**Fig 2.**
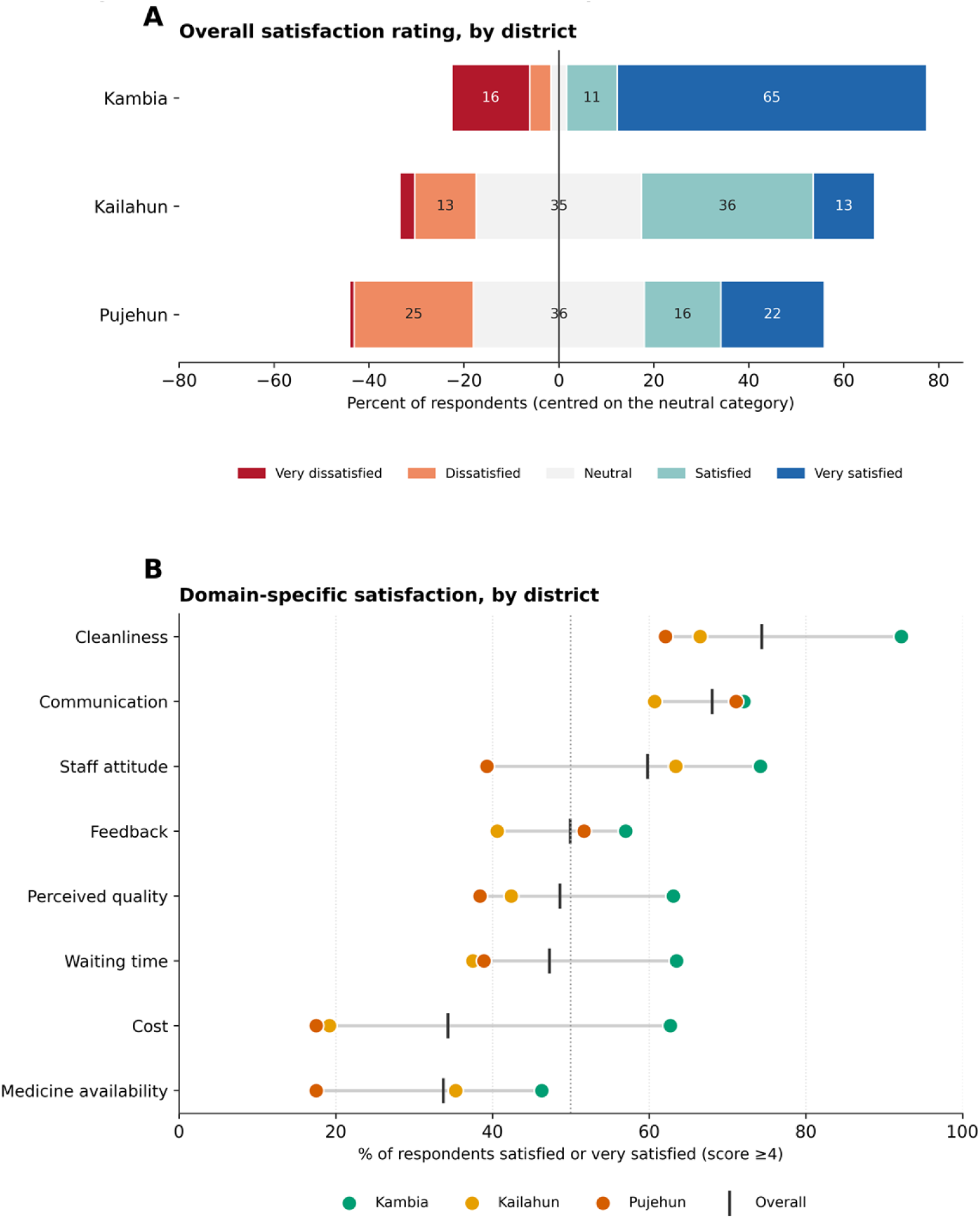
Patient satisfaction with care, by district. (A) Diverging stacked bars centred on the neutral category, showing the full five-point distribution of the single-item overall satisfaction rating in each district. (B) Percentage of respondents satisfied or very satisfied (score ≥4) on each service-experience domain, by district, with the overall value marked (vertical tick); domains are ordered by overall satisfaction.

### Predictors of overall satisfaction

In the multivariable proportional-odds model (n = 676; Table 2, Fig 3A), staff attitude was the strongest independent predictor of higher overall satisfaction: each one-point increase was associated with an AOR of 2.74 (95% CI 2.03–3.70; p<0.001). Waiting-time satisfaction (AOR 1.89 (95% CI 1.39–2.46; p<0.001)) and medicine availability (AOR 1.43 (95% CI 1.16–2.10; p=0.024)) were also independently associated with higher satisfaction, as was visit frequency (AOR 1.14 (95% CI 1.02–1.24; p=0.014) per additional visit). Older age was associated with slightly lower satisfaction (AOR 0.85 (95% CI 0.74–1.00; p=0.028) per 10 years). After adjustment, sex (female vs male, AOR 0.69, 95% CI 0.39–1.03), facility type (hospital vs CHC, AOR 1.20) and travel-time category (AOR 0.91) were not significantly associated with satisfaction.

**Fig 3.**
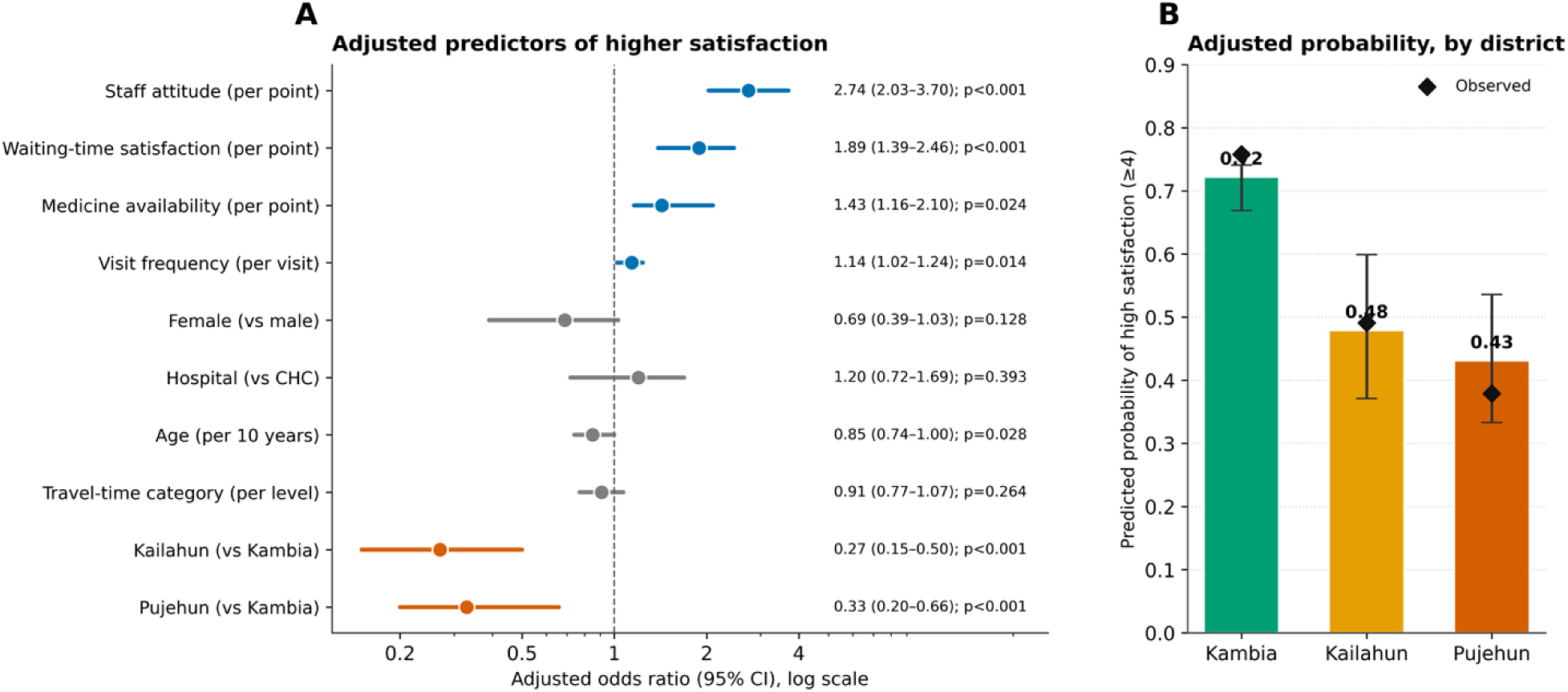
Adjusted predictors and district differences in patient satisfaction. (A) Adjusted odds ratios and 95% confidence intervals from the multivariable proportional-odds ordinal logistic model (n = 676); the dashed line marks the null (OR = 1) and estimates whose 95% CI excludes 1 are shown in colour. (B) Model-predicted probability of high satisfaction (score ≥4) by district with 95% confidence intervals; diamonds show the observed proportions. All confidence intervals are from a cluster bootstrap over the 20 chiefdom clusters.

**Table 2.** Multivariable proportional-odds ordinal logistic regression: adjusted predictors of higher overall patient satisfaction (n = 676)

| Predictor | Adjusted OR | 95% CI | p-value |
| --- | --- | --- | --- |
| Staff attitude (per point) | **2.74** | **2.03–3.70** | **<0.001** |
| Waiting-time satisfaction (per point) | **1.89** | **1.39–2.46** | **<0.001** |
| Medicine availability (per point) | **1.43** | **1.16–2.10** | **0.024** |
| Visit frequency (per visit) | **1.14** | **1.02–1.24** | **0.014** |
| Age (per 10 years) | 0.85 | 0.74–1.00 | 0.028 |
| Female (vs male) | 0.69 | 0.39–1.03 | 0.128 |
| Hospital (vs CHC) | 1.20 | 0.72–1.69 | 0.393 |
| Travel-time category (per level) | 0.91 | 0.77–1.07 | 0.264 |
| Kailahun (vs Kambia) | **0.27** | **0.15–0.50** | **<0.001** |
| Pujehun (vs Kambia) | **0.33** | **0.20–0.66** | **<0.001** |

OR = odds ratio; CI = confidence interval; CHC = community health centre. Estimates are from a single multivariable proportional-odds ordinal logistic model fitted to the five-level overall satisfaction outcome. 95% CIs and p-values were obtained by a cluster bootstrap (1,000 resamples) over the 20 chiefdom clusters. Odds ratios above 1 indicate higher odds of reporting a higher satisfaction category. Reference categories: Kambia (district), community health centre (facility type), male (sex). Continuous service-experience scores (staff attitude, waiting time, medicine availability) are per one-point increase; age is per 10 years; visit frequency is per additional visit; travel time is per category increase. Statistically significant rows (95% CI excluding 1) are shown in bold.

Large district differences persisted after full adjustment. Relative to Kambia, the odds of reporting higher satisfaction were markedly lower in Kailahun (AOR 0.27 (95% CI 0.15–0.50; p<0.001)) and Pujehun (AOR 0.33 (95% CI 0.20–0.66; p<0.001)). The corresponding model-predicted probabilities of high satisfaction were 0.72 (95% CI 0.67–0.74) in Kambia, 0.48 (0.37– 0.60) in Kailahun and 0.43 (0.33–0.54) in Pujehun (Fig 3B).

### Sensitivity analyses and model checks

Findings were robust to model specification. A binary logistic model for high satisfaction (score ≥4) reproduced the direction and significance of all effects (staff attitude AOR 2.17, 95% CI 1.67–3.51; waiting time 1.93, 1.45–3.02; medicine availability 1.71, 1.17–2.47; Kailahun 0.14 and Pujehun 0.11 vs Kambia) (S4 Table). An ordinal model based on the five-domain composite score yielded the same qualitative conclusions. Comparison of cut-point-specific coefficients showed the direction of associations was preserved across all thresholds, supporting the proportional-odds assumption for the principal predictors (S5 Text). Because all analytic respondents had complete outcome and experience data, results were unchanged under complete-case handling.

## Discussion

In this district-comparative survey of 679 recent facility users across three rural border districts of Sierra Leone, patient satisfaction was driven principally by modifiable features of the service encounter, respectful staff behaviour, shorter waits and reliable medicine supply, rather than by fixed demographic or geographic characteristics. Just over half of users (55.2%) were satisfied or very satisfied overall, but this masked a steep district gradient that persisted after adjustment, from a predicted 0.72 probability of high satisfaction in Kambia to 0.43 in Pujehun. The single largest lever was interpersonal: each one-point gain in staff attitude was associated with roughly a two-and-three-quarter-fold increase in the odds of higher satisfaction.

These results are consistent with a recent systematic review of sub-Saharan African studies that identified provider attitude and waiting time as the two most consistent determinants of patient satisfaction [8], and with facility-based studies in Ethiopia and Tanzania that emphasise interpersonal quality and queue management [6,7]. The magnitude of the staff-attitude association in our setting exceeded pooled estimates from urban facilities in Kenya and Ghana, suggesting that interpersonal quality may be especially salient where health-worker density is low and provider–patient power asymmetries are pronounced [9]. The absence of an independent association with travel time contrasts with some reports from the region but is plausible given the widespread use of motorcycles once a decision to seek care has been made [10]; the burden of distance may operate earlier, on the decision to seek care, rather than on the experience of care received. The moderate but robust role of medicine availability underscores that supply-chain reliability is a complementary quality lever, not a peripheral one, particularly given that medicine availability and cost were the lowest-scoring domains in every district.

### Interpretation for the UHC agenda

Sierra Leone’s UHC Roadmap identifies respected, accepted and responsive services as prerequisites for sustained demand and better outcomes [3]. Three operational priorities follow directly from these findings. First, quality-improvement plans should embed structured training and supportive supervision to strengthen respectful, patient-centred communication, the factor most strongly associated with satisfaction and one that requires no capital investment. Second, waiting times should be reduced through workflow redesign, triage and process mapping to relieve bottlenecks in patient flow [11]. Third, routine, facility-level collection and use of patient-experience data, analysed by facility management committees and district health management teams, would institutionalise accountability and support continuous improvement. These priorities align with the National Health Sector Strategic Plan’s goal of improving health-system responsiveness and with the WHO UHC service-coverage framework, which treats patient experience as a dimension of quality [1,10].

### District disparities and equity

The persistence of large district effects after adjustment for individual and service-experience factors indicates that the gap between Kambia and the other districts reflects differences in service delivery and system performance rather than in the populations served. Pujehun, which scored lowest on most domains despite serving the youngest and most predominantly female population, is a clear priority for targeted investment. Treating district as a random effect did not materially change the covariate estimates, reinforcing that the identified levers are relevant within, as well as between, districts.

### Sex, age and the direction of association

After adjustment, sex was not independently associated with overall satisfaction; the descriptive predominance of women in the sample reflects their greater use of facility-based reproductive and child-health services rather than a difference in satisfaction. This is an important correction to a common assumption and should temper interpretations that treat women’s higher facility use as evidence of higher satisfaction. Older respondents reported modestly lower satisfaction, which may reflect higher expectations or a greater burden of chronic need. The positive association between visit frequency and satisfaction should be read cautiously, as satisfied users may simply return more often; longitudinal designs are needed to establish direction.

### Strengths and limitations

Strengths include a probability-based two-stage sampling design, a validated multidimensional instrument with high internal consistency, design-appropriate inference via a cluster bootstrap, and convergent findings across three model specifications. Several limitations warrant caution. First, the cross-sectional design precludes causal inference; associations, including that between visit frequency and satisfaction, may be bidirectional. Second, self-reported satisfaction is susceptible to social-desirability and courtesy bias, which may inflate absolute levels; our focus on relative, within-study comparisons mitigates but does not eliminate this concern. Third, ethical approval for secondary analysis was obtained after data collection, as the survey was initially designed for programme monitoring; the committee judged the use of fully de-identified data with prior consent to be minimal risk, but prospective review would have been preferable. Fourth, the instrument may not fully capture culturally specific notions of respect and dignity. Finally, the survey followed recent HSSEP activities across the participating facilities, so satisfaction may partly reflect a short-term improvement that could attenuate over time. The modest number of clusters (20 chiefdoms) also limits the precision of design-based inference, which we addressed through bootstrapping rather than large-sample approximations.

### Future research

Longitudinal, mixed-methods studies with real-time patient-feedback dashboards could track satisfaction trends and identify high-performing facilities for peer learning. Implementation trials that pair staff-communication coaching with queue-management redesign are needed to test whether improving these modifiable factors increases service utilisation and health outcomes. Qualitative work should examine how expectations shape reported satisfaction, given that self-reported satisfaction appears high relative to the documented burden of poor maternal, newborn and child health outcomes in these districts.

## Conclusions

Patient satisfaction in rural Sierra Leone is shaped principally by modifiable service-delivery factors, respectful staff interactions, reasonable waiting times and reliable medicine supply, rather than by fixed demographic or geographic characteristics. Interventions that strengthen provider–client communication, streamline patient flow and secure essential-medicine availability could deliver rapid, low-cost gains in perceived quality and narrow the substantial district disparities identified here. Embedding routine patient-experience measurement within national and district quality dashboards would reinforce accountability for people-centred care and accelerate equitable progress towards Universal Health Coverage.

## Declarations

### Ethics approval

Sierra Leone Ethics and Scientific Review Committee (SLESRC 002/10/2025). All participants provided informed consent; parental consent and minor assent were obtained for participants under 18 years.

### Data availability

De-identified individual participant data, the data dictionary, the data-collection tool and analytic code will be made available to researchers who submit a methodologically sound proposal, subject to ethical approval and a data-access agreement, from the corresponding author.

### Funding

German Federal Ministry for Economic Cooperation and Development (BMZ), through the GIZ-implemented Health System Strengthening and Epidemic Prevention (HSSEP) programme. The funder had no role in study design, analysis, interpretation, or the decision to publish.

### Competing interests

The authors declare no competing interests.

### Author contributions

AM conceived the study, curated the data, conducted the analysis and drafted the manuscript. BM, SL and OS coordinated fieldwork and supervised data collection. YS, NS and LS contributed to interpretation and critically revised the manuscript. AM and BM verified the underlying data. All authors approved the final version and accept responsibility for the decision to submit.

## Acknowledgements

We thank the District Health Management Teams of Kailahun, Kambia and Pujehun, the enumerators, and the study participants. We acknowledge technical collaboration from Sustainable Health Systems Sierra Leone and Ministry of Health quality-improvement structures.

## Supporting information

S1 Checklist. STROBE checklist for cross-sectional studies.

S2 Text. Sample-size calculation and sampling design.

S3 Code. Analytic code (Python).

S4 Table. Binary logistic sensitivity model for high satisfaction (score ≥4).

S5 Text. Proportional-odds assumption check and model diagnostics.

S6 Table. Data dictionary.

## Notes

### Competing Interest Statement

The authors have declared no competing interest.

### Author Declarations

Sierra Leone Ethics and Scientific Review Committee (SLESRC 002/10/2025). All participants provided informed consent parental consent and minor assent were obtained for participants under 18 years.

